# Artificial Intelligence and Machine Learning in Aneurysmal Subarachnoid Hemorrhage: Future Promises, Perils, and Practicalities

**DOI:** 10.1101/2023.07.18.23292822

**Authors:** Saif Salman, Qiangqiang Gu, Rohan Sharma, Yujia Wei, Benoit Dherin, Sanjana Reddy, Rabih Tawk, W David Freeman

**Author notes:** Correspondence: W David Freeman Mangurian Bldg., Fourth Floor 4500 San Pablo Road Jacksonville, Florida 32224.

## Abstract

**Introduction:** Aneurysmal subarachnoid hemorrhage (SAH) is a subtype of hemorrhagic stroke with thirty-day mortality as high as 40%. Given the expansion of Machine Learning (ML) and Artificial intelligence (AI) methods in health care, SAH patients desperately need an integrated AI system that detects, segments, and supports clinical decisions based on presentation and severity.

**Objectives:** This review aims to synthesize the current state of the art of AI and ML tools for the management of SAH patients alongside providing an up-to-date account of future horizons in patient care.

**Methods:** We performed a systematic review through various databases such as Cochrane Central Register of Controlled Trials, MEDLINE, Scopus, Cochrane Database of Systematic Reviews, and Embase.

**Results:** A total of 507 articles were identified. Following extensive revision, only 21 articles were relevant. Two studies reported improved mortality prediction using Glasgow Coma Scale and biomarkers such as Neutrophil to Lymphocyte Ratio and glucose. One study reported that ffANN is equal to the SAHIT and VASOGRADE scores. One study reported that metabolic biomarkers Ornithine, Symmetric Dimethylarginine, and Dimethylguanidine Valeric acid were associated with poor outcomes. Nine studies reported improved prediction of complications and reduction in latency until intervention using clinical scores and imaging. Four studies reported accurate prediction of aneurysmal rupture based on size, shape, and CNN. One study reported AI-assisted Robotic Transcranial Doppler as a substitute for clinicians.

**Conclusion:** AI/ML technologies possess tremendous potential in accelerating SAH systems-of-care. Keeping abreast of developments is vital in advancing timely interventions for critical diseases.

## 1. Introduction

Subarachnoid hemorrhage (SAH) is a subtype of hemorrhagic stroke ensuing aneurysmal rupture due to modifiable and non-modifiable factors. Consequent sterile bleeding into the tight subarachnoid space carries a high 30-day mortality of 40% ^1^, as 10-25% of patients will die immediately or upon hospital arrival ^2^. Surviving patients will develop a range of short- and long-term complications peaking on days 4 to 14, such as rebleeding, elevated intracranial pressure (ICP), hydrocephalus, seizures, and cognitive dysfunction. Distinctively, delayed cerebral ischemia (DCI) is an important prognostic factor that develops after the first 24 hours as the sterile intracranial bleed will cause inflammation, micro thrombosis, irritation, and vasospasm of the cerebral arteries. Delayed neuronal damage will impair propagation of depolarization waves and cause generalized autonomic instability affecting the heart and other organ systems as a significant cause of morbidity and mortality ^3^. DCI is seen on imaging as cortical infarcts (single in 40% or bilateral in 50%) of cases distal to the rupture site ^4^.

Artificial Intelligence (AI) algorithms were designed to mimic human abilities to solve problems, identify objects, and recognize patterns. Machine learning (ML) is one of the main subfields in AI that automatically learns patterns and relationships based on the provided data. ML has proved useful in various medical disciplines including figures extraction, defining parameters, and predicting outcomes by anticipating associations between various databases. AI/ML doesn’t require the need for pre-programming and follows a multidisciplinary planning- diagnosis–treatment approach in various radiological, surgical, and non-surgical fields surpassing conventional schemes ^5–7^. ML utilizes unique models such as feedforward artificial neural networks (ffANNs), Random Forest (RF), Support Vector Machine (SVM), Logistic Regression (LR), Stacked Convolutional Denoising Auto-Encoders (SCDAE), Principal Component Analysis (PCA), Deep neural networks (DNN), convolutional neural network (CNN) and Multi-Layer Perceptron (MLP) ^8^. Among these models, ffANN is the most utilized to predict case-specific outcomes competing with decisions made by clinicians^9^.

Several studies implemented AI in imaging classification for Penumbra and Large Vessel Occlusion (LVO). This intimates the cruciality of timely intervention in the prognostication of ischemic strokes through a holistic, systematic approach ^10, 11^. Alas, SAH lacks such an approach. Hence, this systematic review aims to redesign the SAH systems-of-care and synthesize the current state of the art of AI/ML tools available in managing aneurysmal SAH through comparison with the current approach.

## 2. METHODS

This systematic review was reported according to the PRISMA guidelines. Two co-authors (WDF and RS) performed this search. We developed the search criteria and assessment through databases such as Cochrane Central Register of Controlled Trials, MEDLINE, Scopus, Cochrane Database of Systematic Reviews, and Embase (1991, 1946, 1823, 2005, 1974 until present), respectively. We directed our literature search into three domains: 1) clinical, biomarkers and laboratory predictors of diagnosis or prognosis; 2) imaging predictors of diagnosis, prognosis and complications; 3) other literature predicting aneurysm rupture, treatment, or modeling.

Neither publication dates nor languages were limited. We created these search strategies by combining standardized and indexed keywords, titles, and abstracts. Further, we included MesH and Embase search terms and keywords such as artificial intelligence, machine learning, subarachnoid hemorrhage, and aneurysmal subarachnoid hemorrhage.

All results were downloaded, and duplicated citations were identified and removed. Co-authors (QG, BD, and SR) further screened the identified articles, including titles and abstracts. We assessed the quality of results based on the Grading of Recommendations Assessment, Development, and Evaluation (GRADE) criteria from Cochrane (Ryan and Hill, 2016)^12^. Unfortunately, we performed a qualitative assessment due to insufficient data required for conducting a meta-analysis. Co-author (WDF) designed Figure 1 using the BioRender® platform, explaining the current approach for SAH management and hypothesizing our future vision.

**Figure 1:**
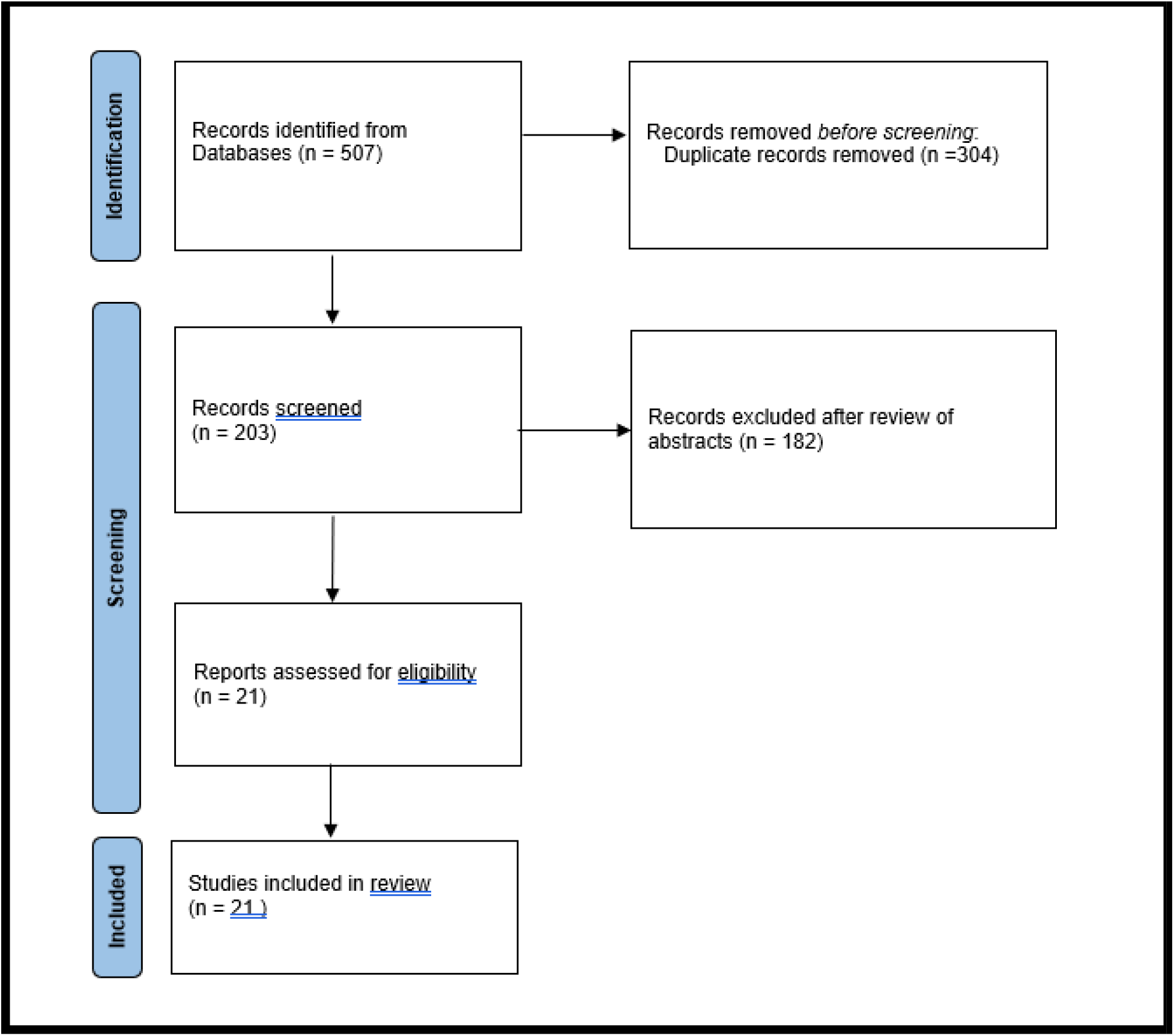
PRISMA diagram detailing the selection process for studies in our systematic review.

## 3. RESULTS

A total of 507 articles were identified. Following the exclusion of duplicates, 203 articles discussing AI/ML and other subtypes of brain hemorrhages remained. Following revision of the abstracts, 182 articles were excluded as they were discussing subtypes of hemorrhagic strokes other than SAH. Hence, 21 articles remained relevant to our study **(Figure 1)**. Upon extensive revision, we used these articles and their bibliography to define our three domains of 1) clinical, biomarkers and laboratory predictors of diagnosis or prognosis; 2) imaging predictors of diagnosis, prognosis, and complications; 3) other literature predicting aneurysm rupture, treatment, or modeling.

### 3.1 Clinical, biomarkers and laboratory predictors of diagnosis or prognosis

Two articles reported improvement in mortality prediction using Glasgow Coma Scale (GCS) and biomarkers such as Neutrophil to Lymphocyte Ratio (NLR) and glucose (Jiewen Deng and Zhaohui He, 2022, Toledo, et al., 2009). In a retrospective study of 1787 patients, Deng et al. reported improvement in mortality prediction in SAH using LR model, compared with GCS, levels of glucose, chloride, bicarbonate, sodium, saturation of oxygen (SPO2), white blood cell count (WBC), temperature, sepsis-related organ failure assessment (SOFA) score, and heparin use. Moreover, Toledo, et al. reported improved prediction outcomes in SAH patients using LR model and comparing them with the World Federation of Neurological Surgeons Committee scale (WFNS), Glasgow Outcome Scale (GOS), and Fisher scale.

A large prospective study of 1226 patients was conducted by Jong et al., where the database included hospital mortality, six months unfavorable modified Rankin Scale (mRS), and incidents of DCI. They found that the feed-forward artificial network (FFAN) equals the Subarachnoid Hemorrhage International Trialists (SAHIT) prediction models and VASOGRADE clinical scoring system in predicting DCI (Jong et al., 2021). Savarraj et al. conducted a retrospective study involving 1032 patients. They compared the performance of LR model with the Fisher scale, age, DCI status, and modified Rankin Scale at discharge and three months. Jude et al. reported that ML models exhibited a significant 36% improvement in the area under the curve (AUC) compared to the LR model when predicting DCI. Furthermore, the AUCs of the ML models were 9% and 18% higher than those of the standard models in predicting discharge and 3-month functional outcomes, respectively (Jude, et al., 2021).

Koch, et al. conducted a study that included 81 CSF samples from patients with SAH, he used an Elastic Net method (an extension of the LR model) and predicted poor outcomes upon correlating Ornithine, Symmetric Dimethylarginine (SDMA), and Dimethylguanidine Valeric acid (DMGV) metabolic biomarkers with the progression of SAH (Koch, et al., 2020). Further, they hypothesized that vasospasm was not the sole factor in DCI development due to variations in levels of nitric oxide (NO), NO synthase, and its precursor arginine. Tanioka, et al. used RF model to analyze (periostin, osteopontin, and galectin-3) multicellular proteins and considered ML in promoting early detection of DCI.

### 3.2 Imaging predictors of diagnosis, prognosis and complications

Eight studies reported improved stratification, accuracy, prediction of complications including DCI, and most importantly, reduction in latency until intervention using total blood volume calculation, and non-contrast CT imaging (NCCT) (Nishi, et al.,2021, Gibson, et al., 2022, Rava, et al. 2021, 2022, Hu, et al., 2022, Ramos, et al., 2019, van der Steen, et al., 2019, Seyam, et al., 2022).

The accuracy of AI in detecting SAH was equal to the accuracy of five neurosurgery specialists to detect SAH based on NCCT scans and surpassed other non-neurosurgical experts (Nishi, et al.). The authors used NCCT images to prepare CNN models for comparison. Hence, suggesting AI as a solution for false negative errors when diagnosing SAH. Gibson, et al. retrospectively studied 46057 patients and used DNN as prediction tools compared to the NCCT imaging to generate two confidence scores (calibrated classifier and Dempster-Shafer) which could reliably detect SAH.

Rava, et al. suggested improved timely intervention as AI could accurately detect SAH and rule out SAH when not present. They conducted a retrospective study, to compare an AI algorithm (Cannon auto Stroke solution)® with NCCT imaging. In a prospective study of 317 patients, utilizing modified Fisher scale, Hunt and Hess clinical score (HHS) and conventional ML models, Ramos, et al. showed that ML can accurately predict DCI and outcome by providing an algorithm that can extract images from NCCT.

Van der Steen, et al. assessed TBV via an automatic quantification algorithm compared to models that used a modified Fisher score to predict DCI and showed that TBV was the sole factor that could accurately predict DCI. Implementing AI-based tools for detecting SAH in a prospective study by Seyam, et al. resulted in 93% accuracy, 87.2% sensitivity, and 97.8% negative predictive value (NPV). Further, a prospective study of 404 patients with SAH showed that ML surpasses the conventional LR methods in predicting DCI and classifying patients with high risk, thus, facilitating interventions (Hu, et al., 2022).

### 3.3 Literature predicting aneurysm rupture, treatment, or modeling

Four studies reported accurate prediction, diagnoses, and evaluation of aneurysmal rupture based on size, shape (Din, et al., 2022, Kim, et al., 2019, 2020, Li, et al., 2022). AI algorithms were suggested to have a promising role in aiding clinicians when diagnosing SAH. However, the evidence is limited to support their adoption in routine clinical care (Din, et al., 2022). Still, implementing AI in assessing rupture probability has been reported to be “feasible” in small-sized aneurysms and surpassing clinicians, using CNN and prospectively testing them in 272 patients (Kim, et al., 2019).

Deploying ML in designing a two-staged hybrid gene selection algorithm and analyzing transcriptomes revealed that (AADAC, LRRCC1, WDR62, RAB32, ANPEP, YIPF1, GZMK, WBP2NL, TOR1B, and PBX1) genes could predict the rupture status of aneurysms and can be used as adjuncts to imaging (Li, et al., 2022). Moreover, Shi, et al., 2020 reported in their review article that using ML algorithms in assessing CTA and MRA scans aid clinicians in managing SAH and predicting outcomes, thus decreasing the clinical workload (Shi, et al., 2020).

A final category regarding AI utilization in Robotics, Esmaeeli, et al., 2020 reported utilizing AI in programming Robotic Transcranial Doppler (TCD), a technique that can surpass manually assessing DCI.

## 4. DISCUSSION

Aneurysmal subarachnoid hemorrhage carries a high risk of mortality and morbidity ^2^. Complications for survivors range from mild brain injury to profound sensory, motor, and cognitive dysfunction^3^. Sterile bleeding into the tight subarachnoid space causes a cascade of inflammation, with consequent complications such as hydrocephalus, vasospasm, and delayed cerebral ischemia. Timely intervention is paramount and can be crucial in determining outcomes^4^.

Artificial Intelligence has entered an era of rapidly progressing algorithms and has received attention universally through prediction-diagnosis-management roles that can surpass counterpart human roles ^6^. ML is the main subtype, where algorithms can be designed and interpreted into clinical, biological, and imaging databases without prior programming through the utilization of unique models such as ffANN most of the time ^7–9^.

For ischemic strokes, a holistic theme to approach patients as they emerge through hospital doors to intervention, often referred to as “Door to Needle” and “Door to Puncture”, has improved outcomes ^10^. This has been reflected in Several studies that were based on utilizing AI in imaging classification for Penumbra and large vessel occlusion (LVO), intimating the cruciality of timely intervention in prognosticating ischemic strokes ^10, 11^.

Unfortunately, the current state of care for patients with SAH lacks a similar streamlined approach. We believe that the average time taken from presentation to imaging is 75 minutes. Additionally, thirty minutes are approximately consumed during the imaging process.

Subsequently, another 45 minutes are dedicated for results interpretation. Moreover, the lack of expert physicians interpreting the results is another major obstacle alongside the time delay since diagnosing SAH can be overlooked if assessed by non-expert physicians ^13^. The future state of care for patients with SAH is based on implementing AI based models to detect SAH within minutes. This can bypass the time consumed during imaging and interpretation. Hence, decreasing the “door to procedure” time and expediting intervention (**Figure 2)**.

**Figure 2:**
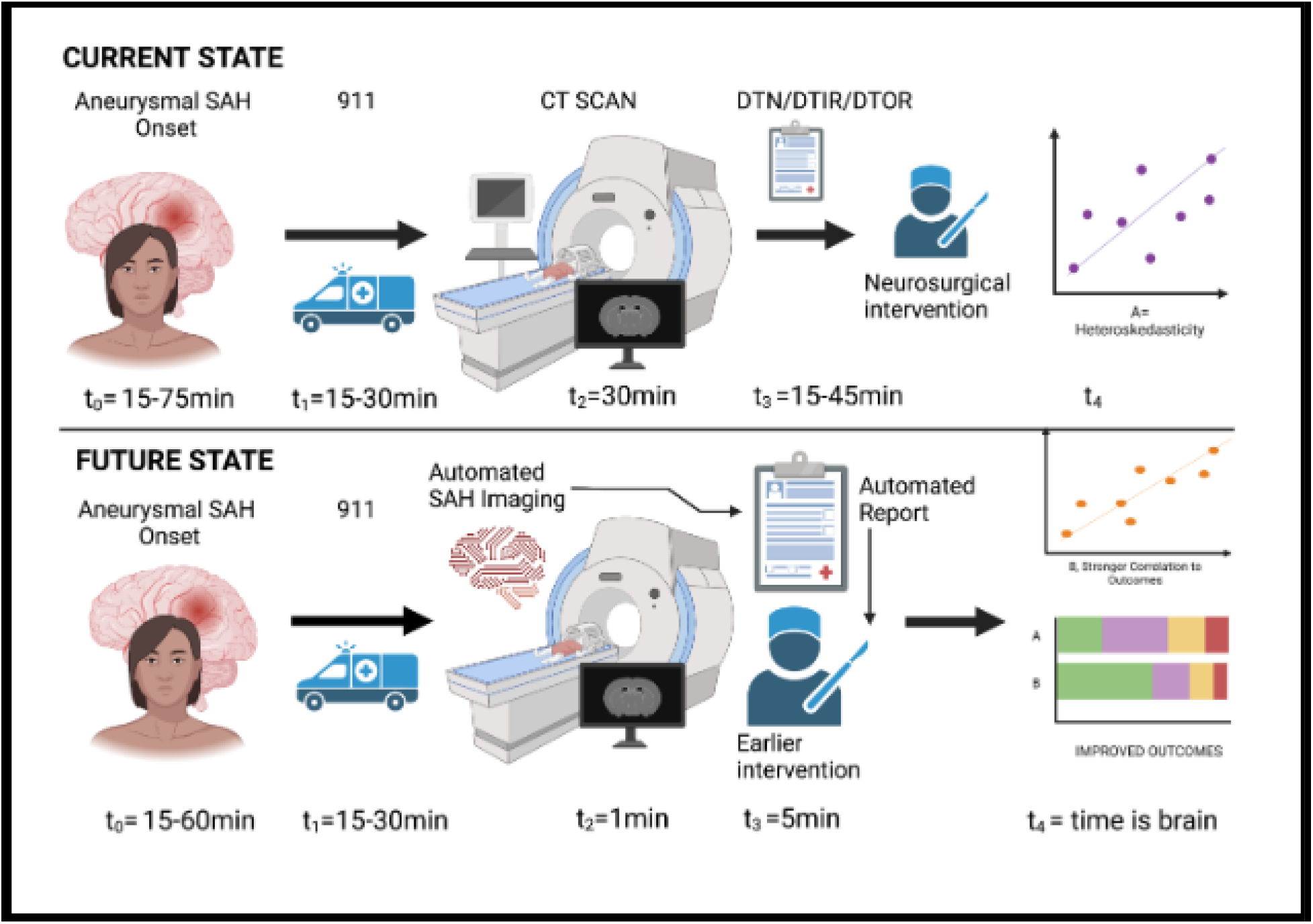
courtesy of Co-author (WDF), hypothesizing the role of AI as a catalyst in reading images and generating reports, Hence, decreasing the imaging to intervention latency.

This is the first systematic review analyzing AI and ML methods for SAH patient care and scientific discovery. Using a streamlined approach, it is now possible to make incredible innovations by translating basic disciplines and promoting patient care. We reviewed studies implementing AI in categories encompassing 1) clinical, biomarkers and laboratory predictors of diagnosis or prognosis; 2) imaging predictors of diagnosis, prognosis, and complications ;3) other literature predicting aneurysm rupture, treatment, or modeling.

### 4.1 Clinical, biomarkers and laboratory predictors of diagnosis or prognosis

Studies that implemented AI/ ML in clinical prediction of diagnosis or prognosis utilizing Fisher, WFNS, SOFA, HHS, GCS, GOS, SAHIT, VASOGRADE clinical scores and studies that implemented AI/ML into biomarkers and laboratory values such as NLR, levels of glucose, chloride, bicarbonate, sodium, SPO2, WBCs, SDMA, DMGV, periostin, osteopontin, and galectin-3 promoted early detection of DCI, mortality prediction, and improvement in general outcome. They further considered AI/ ML utilization to be comparable or superior to our conventional methods ^9, 14–18^. This forebodes a future paradigm of SAH management where AI/ML can aid in diagnosis and outcome predictions for SAH patients leading to a more streamlined approach in their management.

### 4.2 Imaging predictors of diagnosis, prognosis and complications

Studies that implemented AI/ML in the field of imaging predictors reported that AI/ML has the ability to detect SAH at an equivalent level to neurosurgeons while surpassing non-specialists. Furthermore, these studies also showed improved accuracy, predicting complications including DCI, detecting hemorrhagic transformation, and, most importantly, facilitating timely intervention ^8, 13, 19–23^. Hence, we believe that implementing AI and ML in our future protocols can overcome false negative errors in diagnosing SAH when experts are not available, thereby directing appropriate interventions on a case-to-case basis without delay.

### 4.3 Literature predicting aneurysm rupture, treatment, or modeling

Studies that implemented AI and ML to predict rupture of aneurysms can surpass human prediction ability, especially for small-sized aneurysms ^6, 24, 25^. Hence, AI/ML may serve as an adjunct tool to aid physicians in future prevention of SAH. Further studies that implemented AI and ML in programming and robotics show a promising future by surpassing healthcare professional in detecting DCI and other SAH complications^26^.

AI and ML are rapidly progressing, revolutionizing the field of medicine. However, profound studies on AI and ML’s utility in SAH diagnosis and outcomes must be conducted. While ML has various models, with FFANN being the most utilized, the question of whether artificial neural networks or other machine learning techniques can outperform statistical modeling techniques in predicting clinical obstacles still needs a simple answer.

Approaching SAH from the critical facets of imaging, prompt diagnosis, and timely intervention is vital. However, lacking the in-depth discussion of utilizing ML models and biomarkers in different SAH settings and being constrained by the current literature are limitations of this study. Yet, we believe in establishing future frameworks and implementing AI/ML in SAH diagnosis and management as a new streamlined approach similar to those for ischemic strokes. Hence, we can overcome the most crucial peril of delayed diagnosis and management of SAH.

## Conclusion

Various AI and ML methods offer promising tools that can accelerate stroke systems-of-care by providing highly sensitive prehospital and emergency department decision-making systems, automated early SAH detection and prediction of outcomes, improved standardization of intrahospital care as well as providing critical scientific insights into genetics, and mechanistic pathways that can shed light on future neurotherapeutic approaches for this complex disease.

**Table 1:** Summary of the studies included in our systematic review.

| Studies | Type | Clinical Criteria | AI/ML Models | Findings |
| --- | --- | --- | --- | --- |
| Nishi, et al.,2021 | Retrospective study | NCCT | CNN | Accuracy of AI/ML in diagnosing SAH is equal to 5 neurosurgery specialists and higher than nonspecialists |
| Li, et al., 2022 | Retrospective Study | NCCT | LR | AI/ML can be utilized in specific gene analysis alongside imaging to predict rupture of aneurysms |
| Gibson, et al., 2022 | Retrospective Study | NCCT | DNN | AI/ML provides higher accuracy than physicians in classifying and detecting SAH |
| Rava, et al. 2021 | Retrospective Study | NCCT | AutoStroke solution platform | AI/ML provides accurate detection of SAH |
| Jiewen Deng and Zhaohui He, 2022 | Retrospective Study | Biomarkers including glucose levels, GCS,NLR and SOFA score | LR | AI/ML promotes enhanced mortality prediction |
| Koch, et al., 2020 | Retrospective Study | SDMA, DMGV, and ornithine | Elastic Net/LR | AI/ML can anticipate SAH association with poor outcome |
| Hu, et al., 2022 | Multicentric Observational Cohort | NCCT | LR | AI/ML promotes detection, accuracy and intervention for DCI cases |
| Tanioka, et al., 2019 | Prospective Study | Periostin, osteopontin, and galectin-3 | RF | AI/ML promotes accuracy and sensitivity of DCI detection |
| Kim, et al., 2019 | retrospective study | 3D digital subtraction angiography | CNN | AI/ML has accuracy superseding clinicians in predicting rupture risks for small sized aneurysms |
| Van der Steen, et al., 2019 | Prospective Study | Sex, age, medical history of chronic diseases, modified Fisher scale, WFNS score, TBV, aneurysm location and size | LR | AI/ML provides automated calculation of TBV and can predict DCI |
| Toledo, et al., 2009 | Retrospective Study | WFNS, GOS, GCS, and Fisher scales | LR | AI/ML improves accuracy and outcome predictions for SAH |
| Jong et al., 2021 | Prospective Study | Sex, age, medical history of chronic diseases, modified rankin scale, modified Fisher scale, WFNS score, | FFANN | FFAN is equal to the SAHIT and VASOGRADE scores in predicting outcome |
| Esmaeeli, et al., 2020 | Retrospective Study | NCCT and CT angiograms, modified Fisher, hunt and Hess | robotic TCD (Neural Analytics Lucid Robotic System.) ® | Robotic TCD as an alternative to manual TCD in assessing delayed vasospasm |
| Seyam, et al., 2022 | Prospective Study | NCCT | AIdoc medical ® | AI/ML has 93.0% accuracy, 87.2% sensitivity and 97.8% NPV in SAH detection |
| Ramos, et al.,<br>2019 | Prospective Study | Medical history of chronic illness, WFNS, Hunt and Hess and modified Fisher scale | LR, SVM, RFC, MLP, PCA | AI/ML improves DCI prediction |
| Savarraj, et al.,<br>2021 | Retrospective Study | Fisher scale, age, DCI status, modified Rankin Scale at discharge and 3-month | LR | AI/ML Improves detection, and outcome prediction |

## Data Availability

All data produced in the present work are contained in the manuscript

